# Is iron deficiency a major cause of anaemia in Sri Lankan children aged 5-10 years?

**DOI:** 10.1101/2020.04.18.20070359

**Authors:** Renuka Jayatissa, DN Fernando, JM Ranbanda, KH De Silva

## Abstract

**Background:** To assess the iron status among children aged 5–10 years

**Methods:** A cross sectional study in Gampaha district. A sample of 704 children aged 5-10 years resident in a sample of households selected using a two-stage sampling method. A pre tested interviewer administered questionnaire was used to collect information on socio demographic characteristics, housing and sanitation facilities and other relevant variables from the households. Anthropometry was performed on all children aged 5–10 years using standard procedures. Venous blood samples were taken to assess the haemoglobin (Hb) level, levels of Serum Ferritin and C-Reactive Protein.

**Results:** Overall prevalence rate of anaemia, iron deficiency and iron deficiency anaemia in children aged 5-10 years in Gampaha district was 22.7%, 22.1% and 13.5% respectively. The prevalence of iron deficiency among the urban children was significantly higher than that among rural children. The source of water to the households in the households with piped water had a higher prevalence of iron deficiency which was different from the others. The prevalence of iron deficiency among stunted and wasted children was higher though not at a significant level. A substantial proportion of children who were not anaemic were iron deficient.

**Conclusions:** Iron deficiency is an important health problem in this population. Only half of iron deficient children were anaemic and anaemia cannot be solely explained by iron deficiency.

## INTRODUCTION

Iron deficiency (ID) is the most widespread nutritional deficiency as well as the commonest micronutrient deficiency. According to World Health Organization (WHO), 2 billion people – over 30% of the world’s population – are anaemic with about 1 billion suffering from iron deficiency anaemia [1]. WHO estimates that some 800,000 deaths worldwide are attributed to iron deficiency anaemia (IDA) and it remains among 15 leading contributors to the global burden of disease [2]. As measured in disability adjusted life years (DALYs), iron deficiency anaemia (IDA) accounts for 25 million, or 2.4%, of the total [3].

Iron deficiency is defined as a condition in which there are no iron stores that could be mobilized and in which signs of a compromised supply of iron to tissues, including the erythron, are noted. The more severe stages of iron deficiency are associated with anaemia [4].

Iron deficiency can affect all populations and age groups, but the most vulnerable groups are women and children. It can impair cognitive performance at all stages of life [3]. However, relative contribution of ID to childhood anaemia varies widely across populations and age groups [5]. Additional factors, such as infections, haemoglobinopathise are understudied contributors to anaemia in tropical settings [6].

The unmet need for iron in times of rapid growth, during infancy, early childhood results in iron-deficiency and iron deficiency anaemia. In addition, poor dietary iron intake, especially of bio available iron, and helminth infections such as hookworms result in iron deficiency. The high prevalence of ID and IDA in this population might lead to multiple problems, such as impaired cognitive function, reduced school performance and poor reproductive health. These in turn would impact the future quality of human resources [19]. Physical performance is known to be affected by iron deficiency in the absence of anaemia [18].

In Sri Lanka, anaemia has continued to be an important public health problem as shown in many studies and anaemia is commonly used as an indicator of iron deficiency in population-based surveys. However, there is a clear downward trends have been observed for the prevalence of anaemia in primary schoolchildren in Sri Lanka. The prevalence of anaemia among primary schoolchildren was reported as 58%, 21% in year 2001, 12.1% in 2002 and 16.7% in 2004 at national level [7,8,9,10]. There has been little documentation of the prevalence of iron deficiency in primary schoolchildren. A national level study indicated that the prevalence of iron deficiency in children between 6-59 months (n=7400) was 30% and anaemia was reported as 15.1% [11].

Ministry of health has initiated weekly iron supplementation programme for all primary school children in the country through schools. It was important to identify the magnitude of iron deficiency among primary schoolchildren to assess the benefit of such programme.

Hence this study was conducted to assess the anaemia, iron deficiency and iron deficiency anaemia in children aged 5-10 years.

## SUBJECTS AND METHODS

This is a cross sectional community based study among children 5-10 years of age representing one district (Gampaha) of the country out of 25 districts. This district consists of the second highest population (2.3 million) in Sri Lanka and it has better representation of both urban and rural sectors.

The calculated sample size was 710 considering the prevalence estimates of iron deficiency among children as 30% with 95% of confidence interval and 5% error. Design factor was taken as 2.0 and non respondent rate was considered at 10%.

A multi-stage sampling technique was used to identify the sample. During the first stage all public health midwife areas (PHM) in Gampaha district, which is the smallest health unit, consists of 3000 population, was listed out by urban and rural sectors and 16 PHM areas (identified as clusters) from each sector were randomly selected. Altogether 32 clusters were selected. As the second stage, block consists of 100 households, were marked in the map of PHM area and one block was randomly selected. Households in the selected block was listed out and first household to be visited was randomly selected, from their next household in the closest proximity was visited till twenty two children were included from each PHM area. Five teams each comprising of 2 health personnel who had previous experience of participating in nutritional surveys was responsible for data collection. A pre tested interviewer administered questionnaire was administered to obtain date of birth, sex of children, information on housing and sanitary facilities, dietary information, socioeconomic characteristics and other relevant data.

Anthropometric data were collected by specially trained health personnel one in each team. Heights and weights were measured using standard techniques described by the World Health Organization (WHO) [14]. Height was measured by using a stadiometer with the child without footwear and the head held in the Frankfurt horizontal plane. An electronic Seca weighing scale (UNICEF Uni scale) was used to weigh the children with a minimum of clothing without footwear. Accuracy of the weighing scales were checked every morning using standard weights. The weights and heights were recorded to the nearest 0.1kg and 0.1cm respectively. The measurer variation was assessed by duplicating the 10% of measurements by the same measurer and repeating the 10% by the Nutrition Assistant.

Venous blood samples were taken from all children to assess the levels of haemoglobin (Hb), serum ferritin (SF) and C-reactive protein (CRP). Children who had Hb below 10g/dl was assessed for haemoglobinopathies using HPLC method. WHO defined cut-off levels were used to assess the anaemia (Hb<11.5g/dl), iron deficiency (SF<15µg/l). Acute phase of infections were identified when CRP is more than 5.0mg/L as indicated in the assessment kit [15,16].

All fieldwork was completed during the period, May to June 2011. Ethical approval was obtained from the institutional ethical committee. Permission was obtained from the Provincial and district health authorities. Written informed consent was obtained from parents.

Age was calculated from the subject’s date of birth. Height-for-age and BMI-for-age-sex Z scores were calculated by using Anthro 2010 (WHO) software. The WHO cut-off levels were used to estimate prevalence of stunting (height-for-age <-2SD), prevalence of thinness (BMI-for-age-sex <-2SD) [17]. Chi square test and odds ratio was calculated and the level of significance was taken at the level of < 0.05.

## RESULTS

Of the 710 participants, 702 children (98.9%) provided blood samples and included in the analysis. Gender distribution in the sample was 52.7% of boys and 47.3% of girls. Among those studied, 41.9% from the age group of 5-6.9 years, 40% from 7-8.9 years and 18.1% from 9-10.9 years. A high proportion of mothers with secondary school education (71.4%) and a 36.2% belonged to the income category of 20,000 to 31,999 Sri Lankan Rupees. Majority of the households had 4-6 members (83.3%) and 78% obtained drinking water from tap or protected well. Over 90% of households had sanitary toilet, electricity, television, radio and mobile phone (Table 1).

**Table 1:** Characteristics of the study population (n=702)

| Background characteristic | No. (%)* |
| --- | --- |
| Age of the child (years) |  |
| 5.0 – 6.9 | 294 (41.9) |
| 7.0 – 8.9 | 281 (40.0) |
| 9.0 – 10.9 | 127 (18.1) |
| Sex of the child |  |
| Male | 370 (52.7) |
| Female | 332 (47.3) |
| Sector |  |
| Urban | 349 (49.7) |
| Rural | 353 (50.3) |
| Mother's education in years |  |
| 1-5 | 33 (4.7) |
| 6-10 | 501 (71.4) |
| 11-12 | 158 (22.5) |
| Higher | 10 (1.4%) |
| No. of household members |  |
| ≤ 3 | 100 (14.2) |
| 4-6 | 585 (83.3) |
| ≥7 | 17 (2.4) |
| Monthly household income (Rs.) |  |
| <9,000 | 50 (7.2) |
| 9,000-13,999 | 123(17.6) |
| 14,000-19,999 | 183 (26.2) |
| 20,000-31,999 | 253 (36.2) |
| ≥ 32,000 | 90 (12.9) |
| Source of drinking water |  |
| Tap water | 200 (28.6) |
| Protected well (well, tube) | 345 (49.4) |
| Other (canal, rain water, unprotected well) | 154 (22.0) |
| Toilet facilities |  |
| Sanitary | 675 (96.2) |
| Not sanitary (pit latrine) | 27 (3.8) |
| Type of wall of the house |  |
| Brick | 404 (58.6) |
| Cement blocks | 230(33.3) |
| Other (cadjan, mud, wood, hardboard) | 56 (8.1) |
| Type of roof of the house |  |
| Asbestos | 193 (28.0) |
| Tile | 413(59.9) |
| Other (cadjan, corrugated sheet, concrete) | 84 (12.1) |
| Type of floor of the house |  |
| Cement | 525 (76.1) |
| Tile | 129(18.7) |
| Other (mud, wood, cowdung) | 46 (5.2) |
| Presence of household belongings |  |
| Electricity | 654 (97.3) |
| Refrigerator | 465(69.2) |
| Television | 630 (93.8) |
| Radio | 614 (91.4) |
| Mobile phone | 614 (91.4) |

\*Sub totals differ from total number of children studied due to missing values

As shown in table 2, the prevalence of anaemia was 22.6%; 40.9% were moderate degree, 59.1% were mild degree and no severe cases of anaemia (Hb <8g/dl) were detected. Because limited sample volumes were available, serum ferritin was performed in 644 children (90.7% of the sample). Iron deficiency (ID) was 22.1% and iron deficiency anaemia (IDA) was 2.8%. Only 6 cases Thalssaemia was observed providing 0.9% prevalence in this population. The overall prevalence rates of stunting and thinness were 6.3% and 27.7% respectively. Only 6.3% had CRP values of >5 mg/L indicating acute infections.

**Table 2:** Prevalence of nutrition and micronutrient status of the study population.

| **Nutrition and micronutrients** | **% (95%CI\*)** | **No. of children** |
| --- | --- | --- |
| Anaemia | 22.4 (19.7-25.9) | 702 |
| Severe (<8.0g/dl) | 0.0 |  |
| Moderate (8.0-10.9g/dl) | 40.9 |  |
| Mild (11.0-11.4 g/dl) | 59.1 |  |
| Iron deficiency | 23.7 (19.8-26.2) | 644 |
| Iron deficiency anaemia | 3.3 (1.8-4.4) | 654 |
| Median iodine concentration (25<sup>th</sup> -75<sup>th</sup> ) | 100.7 (68.1-162.3)** | 683 |
| Thalassaemia | 0.9 (0.4-1.7) | 697 |
| Acute infections | 6.3 (4.7-8.3) | 702 |
| Thinness | 27.7 (24.5-31.1) | 698 |
| Stunting | 6.3 (4.7-8.4) | 699 |

(\*CI = confidence interval; \*\*25<sup>th</sup> and 75<sup>th</sup> percentiles)

A significantly high percentage of anaemic children used drinking water from the source of unprotected well, canal and rain water (27.7% versus 20.4%; P=0.03); did not possess a refrigerator in the household (37% versus 29%; P=0.04) than non anaemic children. No significant differences were found regarding age, sex, sector, mothers education, number of household members, sex of the household, monthly household income, toilet facilities, type of the household and household belongings. A significantly high percentage of ID children were living in the urban sector (60.5% versus 46.7%; P=0.002), using tap water as a source of drinking water (37.2% versus 26.8%; P=0.02), living in households did not posses television (12.1% versus 5.3%; P=0.006), radio (32.9% versus 22.3%; P=0.008) and refrigerator (37.9% versus 29.0%; P=0.03) than the non ID children. No significant differences were found between ID and non ID children regarding age, sex, mothers education, number of household members, sex of the household, monthly household income, toilet facilities and type of the household (Table 3).

**Table 3.**
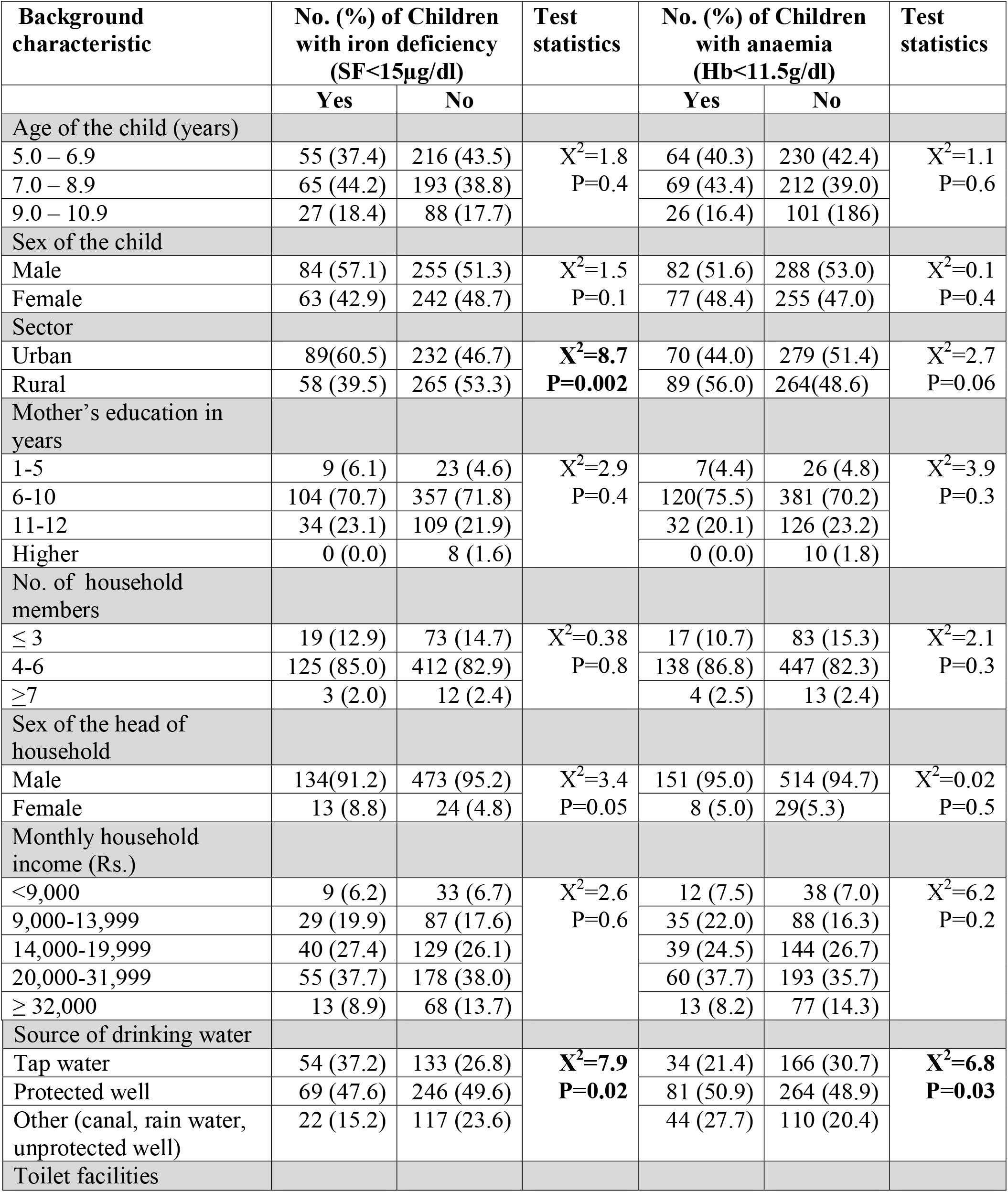
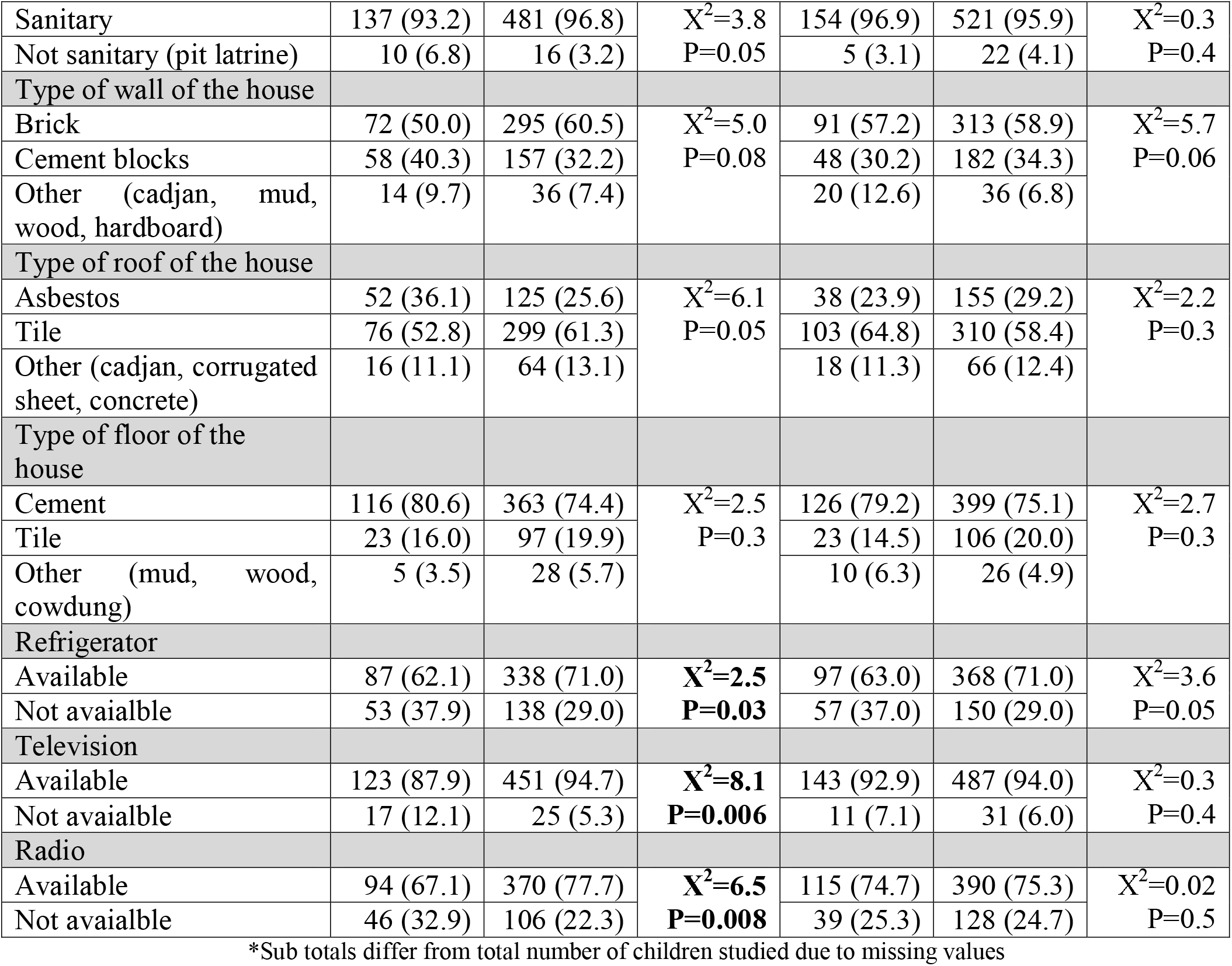
Prevalence of anaemia and iron deficiency among children aged 5-10 years, by socio demographic and socio economic characteristics (N=702*)

| Background characteristic | No. (%) of Children with iron deficiency (SF<15µg/dl) |  | Test statistics | No. (%) of Children with anaemia (Hb<11.5g/dl) |  | Test statistics |
| --- | --- | --- | --- | --- | --- | --- |
|  | Yes | No |  | Yes | No |  |
| Age of the child (years) |  |  |  |  |  |  |
| 5.0 – 6.9 | 55 (37.4) | 216 (43.5) | X<sup>2</sup>=1.8<br>P=0.4 | 64 (40.3) | 230 (42.4) | X<sup>2</sup>=1.1<br>P=0.6 |
| 7.0 – 8.9 | 65 (44.2) | 193 (38.8) |  | 69 (43.4) | 212 (39.0) |  |
| 9.0 – 10.9 | 27 (18.4) | 88 (17.7) |  | 26 (16.4) | 101 (18.6) |  |
| Sex of the child |  |  |  |  |  |  |
| Male | 84 (57.1) | 255 (51.3) | X<sup>2</sup>=1.5<br>P=0.1 | 82 (51.6) | 288 (53.0) | X<sup>2</sup>=0.1<br>P=0.4 |
| Female | 63 (42.9) | 242 (48.7) |  | 77 (48.4) | 255 (47.0) |  |
| Sector |  |  |  |  |  |  |
| Urban | 89(60.5) | 232 (46.7) | X<sup>2</sup>=8.7<br>P=0.002 | 70 (44.0) | 279 (51.4) | X<sup>2</sup>=2.7<br>P=0.06 |
| Rural | 58 (39.5) | 265 (53.3) |  | 89 (56.0) | 264(48.6) |  |
| Mother's education in years |  |  |  |  |  |  |
| 1-5 | 9 (6.1) | 23 (4.6) | X<sup>2</sup>=2.9<br>P=0.4 | 7(4.4) | 26 (4.8) | X<sup>2</sup>=3.9<br>P=0.3 |
| 6-10 | 104 (70.7) | 357 (71.8) |  | 120(75.5) | 381 (70.2) |  |
| 11-12 | 34 (23.1) | 109 (21.9) |  | 32 (20.1) | 126 (23.2) |  |
| Higher | 0 (0.0) | 8 (1.6) |  | 0 (0.0) | 10 (1.8) |  |
| No. of household members |  |  |  |  |  |  |
| ≤ 3 | 19 (12.9) | 73 (14.7) | X<sup>2</sup>=0.38<br>P=0.8 | 17 (10.7) | 83 (15.3) | X<sup>2</sup>=2.1<br>P=0.3 |
| 4-6 | 125 (85.0) | 412 (82.9) |  | 138 (86.8) | 447 (82.3) |  |
| ≥7 | 3 (2.0) | 12 (2.4) |  | 4 (2.5) | 13 (2.4) |  |
| Sex of the head of household |  |  |  |  |  |  |
| Male | 134(91.2) | 473 (95.2) | X<sup>2</sup>=3.4<br>P=0.05 | 151 (95.0) | 514 (94.7) | X<sup>2</sup>=0.02<br>P=0.5 |
| Female | 13 (8.8) | 24 (4.8) |  | 8 (5.0) | 29(5.3) |  |
| Monthly household income (Rs.) |  |  |  |  |  |  |
| <9,000 | 9 (6.2) | 33 (6.7) | X<sup>2</sup>=2.6<br>P=0.6 | 12 (7.5) | 38 (7.0) | X<sup>2</sup>=6.2<br>P=0.2 |
| 9,000-13,999 | 29 (19.9) | 87 (17.6) |  | 35 (22.0) | 88 (16.3) |  |
| 14,000-19,999 | 40 (27.4) | 129 (26.1) |  | 39 (24.5) | 144 (26.7) |  |
| 20,000-31,999 | 55 (37.7) | 178 (38.0) |  | 60 (37.7) | 193 (35.7) |  |
| ≥ 32,000 | 13 (8.9) | 68 (13.7) |  | 13 (8.2) | 77 (14.3) |  |
| Source of drinking water |  |  |  |  |  |  |
| Tap water | 54 (37.2) | 133 (26.8) | X<sup>2</sup>=7.9<br>P=0.02 | 34 (21.4) | 166 (30.7) | X<sup>2</sup>=6.8<br>P=0.03 |
| Protected well | 69 (47.6) | 246 (49.6) |  | 81 (50.9) | 264 (48.9) |  |
| Other (canal, rain water, unprotected well) | 22 (15.2) | 117 (23.6) |  | 44 (27.7) | 110 (20.4) |  |
| Toilet facilities |  |  |  |  |  |  |
| Sanitary | 137 (93.2) | 481 (96.8) | $X^2=3.8$<br>$P=0.05$ | 154 (96.9) | 521 (95.9) | $X^2=0.3$<br>$P=0.4$ |
| Not sanitary (pit latrine) | 10 (6.8) | 16 (3.2) |  | 5 (3.1) | 22 (4.1) |  |
| Type of wall of the house |  |  |  |  |  |  |
| Brick | 72 (50.0) | 295 (60.5) | $X^2=5.0$<br>$P=0.08$ | 91 (57.2) | 313 (58.9) | $X^2=5.7$<br>$P=0.06$ |
| Cement blocks | 58 (40.3) | 157 (32.2) |  | 48 (30.2) | 182 (34.3) |  |
| Other (cadjan, mud, wood, hardboard) | 14 (9.7) | 36 (7.4) |  | 20 (12.6) | 36 (6.8) |  |
| Type of roof of the house |  |  |  |  |  |  |
| Asbestos | 52 (36.1) | 125 (25.6) | $X^2=6.1$<br>$P=0.05$ | 38 (23.9) | 155 (29.2) | $X^2=2.2$<br>$P=0.3$ |
| Tile | 76 (52.8) | 299 (61.3) |  | 103 (64.8) | 310 (58.4) |  |
| Other (cadjan, corrugated sheet, concrete) | 16 (11.1) | 64 (13.1) |  | 18 (11.3) | 66 (12.4) |  |
| Type of floor of the house |  |  |  |  |  |  |
| Cement | 116 (80.6) | 363 (74.4) | $X^2=2.5$<br>$P=0.3$ | 126 (79.2) | 399 (75.1) | $X^2=2.7$<br>$P=0.3$ |
| Tile | 23 (16.0) | 97 (19.9) |  | 23 (14.5) | 106 (20.0) |  |
| Other (mud, wood, cowdung) | 5 (3.5) | 28 (5.7) |  | 10 (6.3) | 26 (4.9) |  |
| Refrigerator |  |  |  |  |  |  |
| Available | 87 (62.1) | 338 (71.0) | $X^2=2.5$<br>$P=0.03$ | 97 (63.0) | 368 (71.0) | $X^2=3.6$<br>$P=0.05$ |
| Not available | 53 (37.9) | 138 (29.0) |  | 57 (37.0) | 150 (29.0) |  |
| Television |  |  |  |  |  |  |
| Available | 123 (87.9) | 451 (94.7) | $X^2=8.1$<br>$P=0.006$ | 143 (92.9) | 487 (94.0) | $X^2=0.3$<br>$P=0.4$ |
| Not available | 17 (12.1) | 25 (5.3) |  | 11 (7.1) | 31 (6.0) |  |
| Radio |  |  |  |  |  |  |
| Available | 94 (67.1) | 370 (77.7) | $X^2=6.5$<br>$P=0.008$ | 115 (74.7) | 390 (75.3) | $X^2=0.02$<br>$P=0.5$ |
| Not available | 46 (32.9) | 106 (22.3) |  | 39 (25.3) | 128 (24.7) |  |

\*Sub totals differ from total number of children studied due to missing values

Regarding food habits as shown in Table 4 anaemic children do not significantly differ from non anaemic children except consuming fish weekly (73.0% versus 59.5%; P=0.009), tubers daily (25.2% versus 14.7%; P=0.009). ID children do not significantly differ from non ID children except eating fruits after main meals (48.3% versus 37.8%; P=0.02) and fish daily (38.1% versus 24.1%; P=0.002).

**Table 4:**
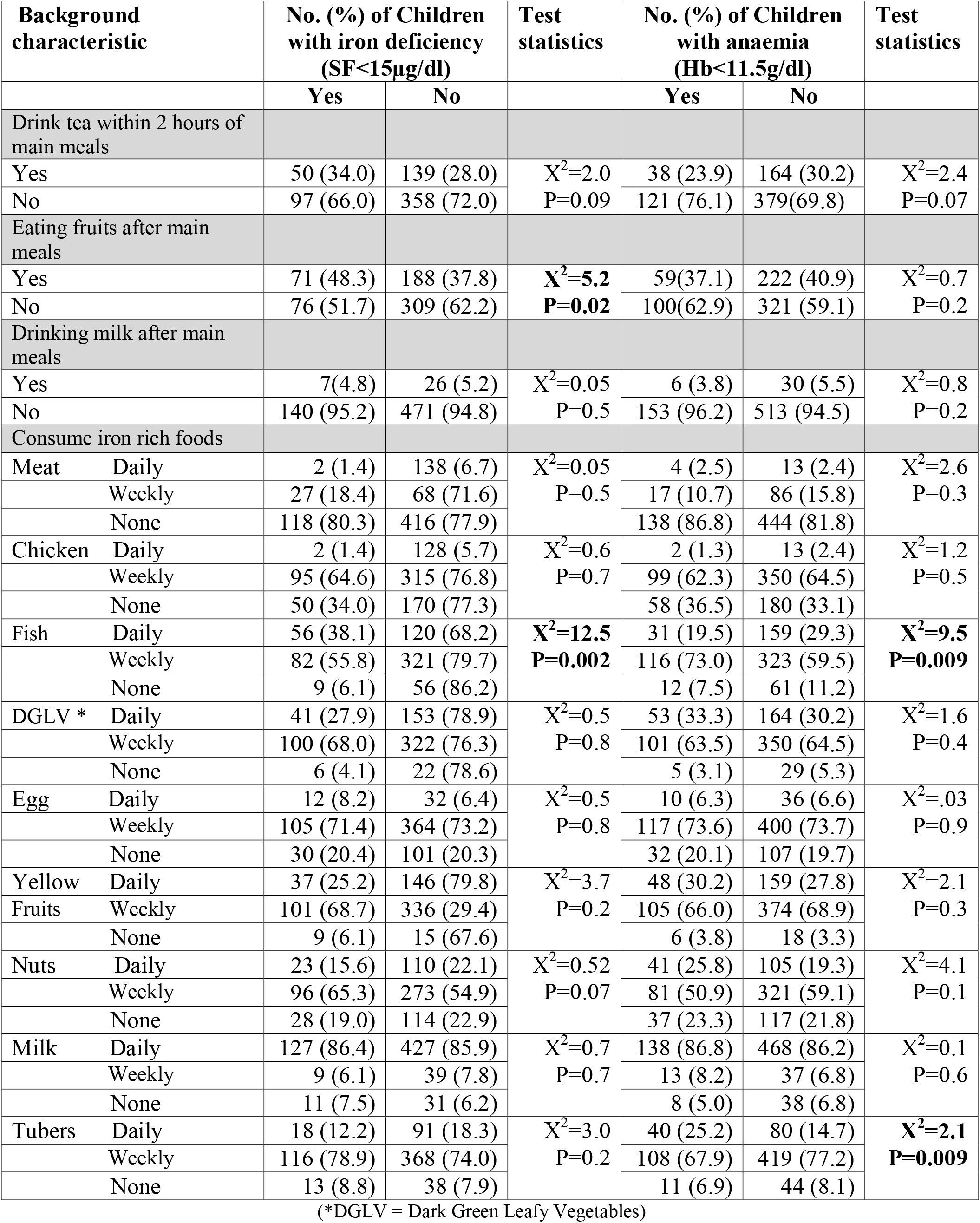
Prevalence of anaemia and ID in relation to selected dietary practices of children.

A total of 14.3% of all ID children were anaemic and 85.7%, whereas the 85.7% of ID children were non anaemic. As shown in table 5, ID children were significantly stunted (10.1% versus 5.2%; P=0.02) and non anaemic (14.3% versus 24.5%; P=0.005) than non ID children. Conversely, anaemic children were significantly less stunted than non anaemic children (14.3% versus 24.5%; P=0.005). Among the anaemic children acute infections were higher than non anaemic children. However this observation was not significant (8.2% versus 5.7%; P=0.2).

**Table 5.** Prevalence of anaema and iron deficiency in relation to other nutritional status indicators.

| Background characteristic | No. (%) of Children with iron deficiency (SF<15µg/dl) |  | Test statistics | No. (%) of Children with anaemia (Hb<11.5g/dl) |  | Test statistics |
| --- | --- | --- | --- | --- | --- | --- |
|  | Yes | No |  | Yes | No |  |
| Thinness |  |  |  |  |  |  |
| Yes | 53 (33.3) | 140 (26.0) | $X^2=3.5$<br>P=0.2 | 11 (7.5) | 29 (5.9) | $X^2=0.5$<br>P=0.3 |
| No | 106 (66.6) | 393 (74.0) |  | 135 (92.5) | 466 (94.1) |  |
| Stunting |  |  |  |  |  |  |
| Yes | 16 (10.1) | 28 (5.2) | $X^2=4.9$<br>**P=0.02** | 21 (14.3) | 122 (24.5) | $X^2=6.9$<br>**P=0.005** |
| No | 143 (89.9) | 512 (94.8) |  | 126 (85.7) | 375 (75.5) |  |
| Anaemia (Hb<11.5g/dl) |  |  |  |  |  |  |
| Yes | 21 (14.3) | 122 (24.5) | $X^2=6.9$<br>**P=0.005** | 21 (14.7) | 126 (25.1) | $X^2=6.9$<br>**P=0.005** |
| No | 126 (85.7) | 375 (75.5) |  | 122 (85.3) | 375 (74.9) |  |
| Acute infections (CRP>5.0) |  |  |  |  |  |  |
| Yes | | | | 13 (8.2) | 31 (5.7) | $X^2=1.3$<br>P=0.2 |
| No |  |  |  | 146 (91.8) | 512 (94.3) |  |
| Mean HAZ (SD) | -0.69<br>(1.1) | -0.62<br>(1.1) | F=0.7<br>P=0.39 | -0.76<br>(1.03) | -0.59<br>(1.1) | **F=4.0**<br>**P=0.04** |
| Mean BAZ (SD) | -1.28<br>(1.2) | -1.27<br>(1.1) | F=0.003<br>P=0.96 | -1.42<br>(1.1) | -1.22<br>(1.19) | F=3.4<br>P=0.06 |

## Discussion

Our study revealed that the overall prevalence rate of anaemia, ID and IDA in children aged 5-10 years in Gampaha district was 22.6%, 22.8% and 2.8% respectively. This highlights that iron deficiency is not the major cause of anaemia in this population. Anaemia in the Sri Lankan children studied is a moderate public health problem (between 20-39.9%) as classified by the World health organization (WHO) [4]. Anaemia remained less frequent than in comparable population groups in Thailand (22.6% versus 31%), but higher than the rates reported by previous studies conducted in Sri Lanka [5,7,8,9,10].

It is generally assumed that worldwide “at least half the anaemia is due to nutritional iron deficiency” and that the prevalence of iron deficiency will be about 2.5 times that of IDA [18]. However, in this study the prevalence of anaemia and iron deficiency are almost the same and iron deficiency is 8 times that of iron deficiency anaemia. Recent reviews revealed that there are considerable variations in both of these ratios depending on the age and sex of the people being studied, the region of the world in which they live, and the prevalence rates of other causes of anaemia. The prevalence of anaemia alone can therefore give only a very rough estimate of the likely prevalence of iron deficiency anaemia [18].

This study indicates 25.1% of those who were not anaemic being iron deficient and 85.3% of those who were anaemic were not iron deficient highlighting the importance of investigating the other possible causes for anaemia. Similar findings were observed among schoolchildren in Thailand [5].

Studies and reviews have demonstrated that the infections play a central role in the pathogenesis of anaemia[13]. In the present study population the prevalence of acute infections (CRP>5mg/L) were reported as 6.3%; 8.2% of anaemic children had infections and 91.8% had no acute infections suggesting acute infections is not a major predictor of anaemia in this population. Previous studies have demonstrated that malaria as a strong predictors of anaemia. However, malaria infections are uncommon in this part of Sri Lanka. ID was calculated excluding the children with CRP>5mg/L. Thalassaemia is not a major cause of anaemia in this population; only 9 cases were found in this.

In this study, anaemia and iron deficiency was not predictors by socioeconomic factors: family income, number of family members, mother’s education except source of drinking water, sector and possession of television and fridge. However, some studies have shown a high prevalence of IDA in families with lower education levels and a high number of family members [19, 20, 21]. Children living in urban areas had a higher chance of developing ID indicating a need to collect in depth information on their dietary practices and other factors that could lead to iron deficiency.

The dietary pattern reflects daily consumption of fish and eating fruits after main meals were significantly associated with ID, may be due to not using iron rich fish and vitamin C rich food. Conversely anaemia was significantly associated with children consuming tubers daily and eating fish weekly. Assessment of variety of food and dose response effects was not studied in this study.

Previous studies have demonstrated that the role of geohelminth infections on anaemia was very high, but the overall prevalence of hookworm infestation in Sri Lanka was reported as 1.2% [10]. Impact of geohelminth infection was not studied in this population and it is better to explore it further.

The unmet need for iron in times of rapid growth, during infancy, early childhood results in iron-deficiency and iron deficiency anaemia. In addition, poor dietary iron intake, especially of bio available iron, and helminth infections such as hookworms result in iron deficiency. The high prevalence of ID and IDA in this population might lead to multiple problems, such as impaired cognitive function, reduced school performance and poor reproductive health. These in turn would impact the future quality of human resources [19]. Physical performance is known to be affected by iron deficiency in the absence of anaemia [18].

## Conclusion

The results suggest that anaemia and ID are an important public health problems in this population and anaemia cannot be solely explained by iron deficiency. Strategies to control anaemia based exclusively on iron supplementation are likely to have only a limited impact on the overall prevalence of anaemia. Further studies are needed to identify the aeitiology of anaemia in this population. The prevalence of iron deficiency cannot be predicted by the prevalence of anaemia and it is necessary to use specific iron indicators to identify iron deficiency to prevent functional consequences.

## Data Availability

With the corresponding author

## ACKNOWLEDGEMENTS

This study was supported by the Micronutrient Initiatives India and coordinated by ICCIDD Sri Lanka. The authors thank Dr. Venkatash Mannar, Mr. Mahinda Gunawardana, Mr. Mahinda Wijesundara for assistance and Dr. Upul Senarath and Indika Siriwardana for statistical support. Thanks for RDHS and health staff of Gampaha district and all the staff of Department of Nutrition for supporting the data collection.

## REFERENCES

1. World Health Organisation (WHO). Nutrition. Geneva: www.who.int/nutrition/en: WHO, 2007.

2. World Health Organisation (WHO). World Health Report 2002: reducing risks, promoting healthy life: overview. Geneva: WHO, 2002.

3. Gleason G and Scrimshaw N. An overview of the functional significance of iron deficiency. Nutritional anaemia. Sight and Life, 2007.

4. World Health Organisation (WHO). Iron Deficiency Anaemia: Assessment, Prevention and Control - A guide for programme managers. Geneva: WHO, 2001.

5. Thurlow RA, Winichagoon P, Green T, Wasantwisut W, Pongcharoen T, Bailey KB, Gibson RS. Only a small proportion of anemia in northeast Thai schoolchildren is associated with iron deficiency. Am J Clin Nutr. 2005; 82: 380–387.

6. Cardoso MA, Scopel KK, Muniz PT, Villamor E, Ferreira MU (2012) Underlying Factors Associated with Anemia in Amazonian Children: A Population-Based, Cross-Sectional Study. PLoS ONE 7(5): e36341. doi:10.1371/journal.pone.0036341

7. Piyasena C, Mahamithawa S. Anaemia status in Sri Lanka. Medical Research Institute, 2001.

8. Mudalige R, Nestel P. Prevalence of Anaemia in Sri Lanka. The Ceylon Journal of Medical Science. 1996; 29: 9–16.

9. Jayatissa R, Ranbanda JM. Nutritional status in primary schoolchildren. Medical Research Institute, 2004. Available at: http://www.mri.gov.lk/nutrition/research and publications/2004. Accessed 12 November 2012.

10. Pathmeswaran A, Jayatissa R, Samarasinghe S, Fernando A, De Silva RP, Thattil R O, De Silva NR, ‘Health status of primary school children in Sri Lanka’ Ceylon Medical Journal, 2006; vol. 50, no. 2 (June), pp: 46–50.

11. Tudawe M, Wijesiriwardena I, Peiris M, Dhammika L. Etiology of Anaemia in Sri Lanka. Ministry of Health, Colombo; 2005.

12. Cook JD (2005) Diagnosis and management of iron-deficiency anaemia. Best Pract Res Clin Haematol 18: 319–332.

13. Stoltzfus RJ, Chwaya HM, Montresor A, Albonico M, Savioli L, et al. (2000) Malaria, hookworms and recent fever are related to anemia and iron status indicators in 0- to 5-y old Zanzibari children and these relationships change with age. J Nutr 130: 1724–1733.

14. World Health Organisation (WHO). Physical status: the use and interpretation of anthropometry. WHO Technical Report series 854. Geneva,1995.

15. WHO. Haemoglobin concentrations for the diagnosis of anaemia and assessment of severity. Vitamin and Mineral Nutrition Information System. Geneva, World Health Organization, 2011 (WHO/NMH/NHD/MNM/11.1) (http://www.who.int/vmnis/indicators/haemoglobin.pdf, accessed [15/10/2012]).

16. WHO. Serum ferritin concentrations for the assessment of iron status and iron deficiency in populations. Vitamin and Mineral Nutrition Information System. Geneva, World Health Organization, 2011 (WHO/NMH/NHD/MNM/11.2). (http://www.who.int/vmnis/indicators/serum_ferritin.pdf, accessed [15/10/2012]).

17. De Onis M, Onyango AW, Borghi E, Siyam A, Nishida C, Siekmann J. Development of a WHO growth reference for school-aged children and adolescents. Bulletin of the World Health Organization 2007;85:660–7.

18. Joint World Health Organization/Centers for Disease Control and Prevention. Technical Consultation on the Assessment of Iron Status at the Population Level. Geneva, Switzerland;2004

19. Mamdooh A. Gari, Prevalence of Iron Deficiency Anemia among Female Elementary School Children in Northern Jeddah, Saudi Arabia, Journal of King Abdulaziz University: Medical Science, 2008; Vol. 15 No. 1, pp: 63–75.

20. Mohamed El Hioui, Ahmed Omar Touhami Ahami, Youssef Aboussaleh, Stephane Rusinek, KhalidDik and Abdelkader Soualem. Iron Deficiency and Anaemia in Rural School Children in a Coastal Area of Morocco. Pakistan Journal of Nutrition, 2008; 7 (3): 400–403.

21. EL Hioui M1, Ahami A.O.T, Aboussaleh Y, Rusinek S, Dik K, Soualem A, Azzaoui F-Z, Loutfi H & Elqaj M. Risk Factors of Anaemia Among Rural School Children in Kenitra, Morocco East African Journal of Public Health,2008; Volume 5, Number 2(August): 62–66.

